## Supplementary material for "Effective vaccine allocation strategies, balancing economy with infection control against COVID-19 in Japan": S1_Table

| *R*_0_ | *E* (*%*) | Vaccine strategy | *T*_L_ (day) | *L* (%) | Infected | Deaths |
| --- | --- | --- | --- | --- | --- | --- |
| 1.1 | 1 | Young-old-middle | 10 | 48.67 | 74827 | 1199 |
| 1.1 | 4 | Young-middle-old | 20 | 97.33 | 14530 | 490 |
| 1.1 | 7 | Old-young-middle | 34 | 99.99 | 11713 | 418 |
| 1.1 | 10 | Old-other | 48 | 99.99 | 10094 | 415 |
| 1.3 | 1 | Young-old-middle | 11 | 44.24 | 173972 | 2501 |
| 1.3 | 4 | Young-other | 23 | 84.64 | 20395 | 566 |
| 1.3 | 7 | Young-middle-old | 34 | 100.00 | 10593 | 442 |
| 1.3 | 10 | Old-young-middle | 48 | 100.00 | 11993 | 420 |
| 1.5 | 1 | Young-other | 13 | 37.44 | 495815 | 7110 |
| 1.5 | 4 | Young-old-middle | 27 | 72.10 | 36034 | 782 |
| 1.5 | 7 | Young-old-middle | 34 | 100.00 | 11369 | 452 |
| 1.5 | 10 | Equal | 49 | 99.32 | 10939 | 437 |
| 1.7 | 1 | Young-old-middle | 13 | 37.44 | 1646979 | 25089 |
| 1.7 | 4 | Young-old-middle | 26 | 74.76 | 78788 | 1459 |
| 1.7 | 7 | Young-other | 35 | 97.33 | 14131 | 493 |
| 1.7 | 10 | Young-old-middle | 48 | 99.94 | 10850 | 444 |
| 1.9 | 1 | Young-old-middle | 14 | 34.75 | 5650738 | 92210 |
| 1.9 | 4 | Young-old-middle | 30 | 64.89 | 220064 | 3858 |
| 1.9 | 7 | Young-old-middle | 35 | 96.99 | 18883 | 574 |
| 1.9 | 10 | Young-old-middle | 48 | 100.00 | 11265 | 449 |
