## Supplementary material for "Effective vaccine allocation strategies, balancing economy with infection control against COVID-19 in Japan": S2_Table

| *R*_0_ | *E* (*%*) | Vaccine strategy | *T*_L_ | *L* | Infected | Deaths |
| --- | --- | --- | --- | --- | --- | --- |
| 1.1 | 1 | Old-young-middle | 12 | 40.55 | 424789 | 1599 |
| 1.1 | 4 | Old-young-middle | 22 | 88.48 | 41692 | 503 |
| 1.1 | 7 | Old-young-middle | 34 | 99.99 | 11713 | 418 |
| 1.1 | 10 | Old-other | 48 | 99.99 | 10094 | 415 |
| 1.3 | 1 | Old-young-middle | 18 | 27.04 | 6153990 | 10411 |
| 1.3 | 4 | Old-young-middle | 26 | 74.87 | 425977 | 1084 |
| 1.3 | 7 | Old-young-middle | 35 | 97.33 | 41322 | 467 |
| 1.3 | 10 | Old-young-middle | 48 | 100 | 11993 | 420 |
| 1.5 | 1 | Old-middle-young | 20 | 24.33 | 36143189 | 43426 |
| 1.5 | 4 | Old-young-middle | 51 | 38.17 | 6561019 | 9742 |
| 1.5 | 7 | Old-young-middle | 52 | 65.51 | 357155 | 986 |
| 1.5 | 10 | Old-young-middle | 51 | 95.42 | 31793 | 456 |
| 1.7 | 1 | Old-middle-young | 18 | 27.03 | 49469117 | 97079 |
| 1.7 | 4 | Old-middle-young | 52 | 37.44 | 40811860 | 34115 |
| 1.7 | 7 | Old-young-middle | 75 | 45.42 | 5304065 | 8333 |
| 1.7 | 10 | Old-young-middle | 68 | 71.51 | 198849 | 817 |
| 1.9 | 1 | Old-middle-young | 24 | 20.28 | 58420160 | 179593 |
| 1.9 | 4 | Old-middle-young | 50 | 38.93 | 51880645 | 60060 |
| 1.9 | 7 | Old-middle-young | 65 | 52.41 | 43069323 | 29179 |
| 1.9 | 10 | Old-young-middle | 79 | 61.53 | 2971898 | 5152 |
