## Supplementary figures and images for "Effective vaccine allocation strategies, balancing economy with infection control against COVID-19 in Japan"

### S3_Fig

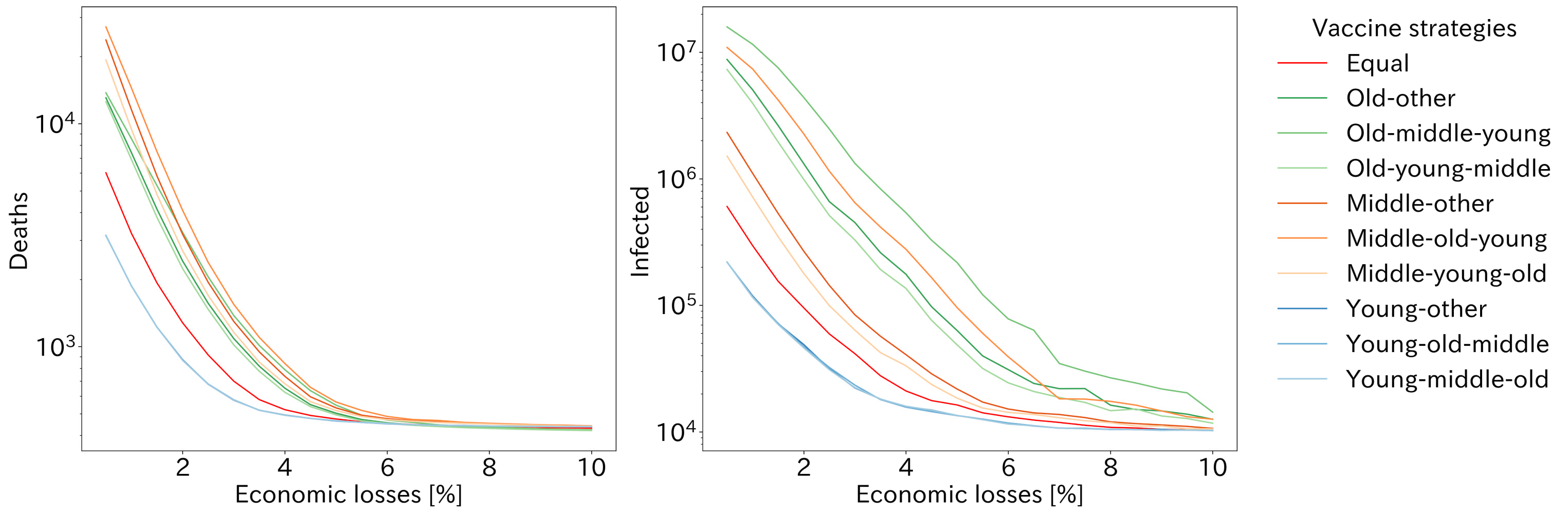

### S4_Fig

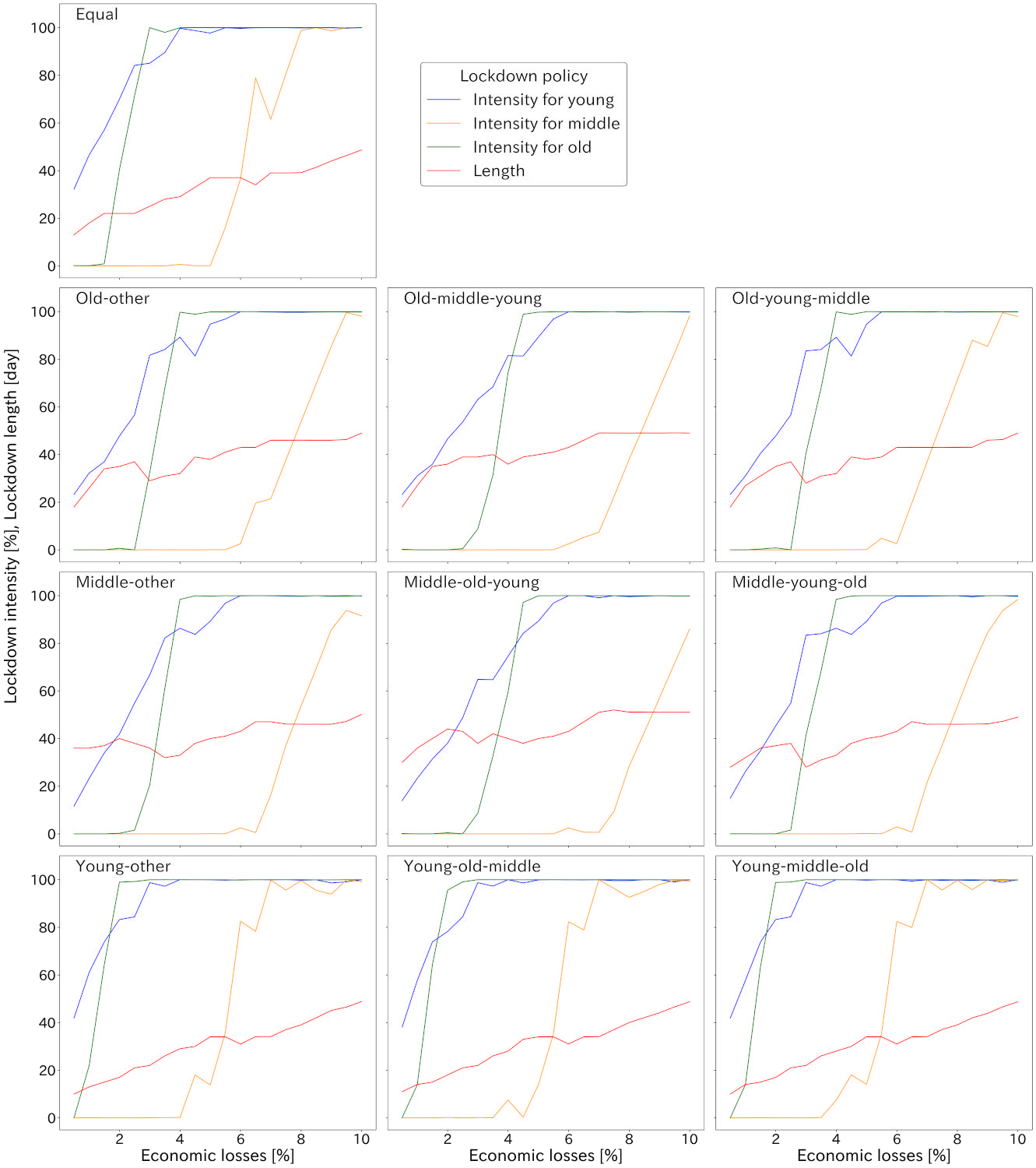
